# The Burr distribution as a model for the delay between key events in an individual’s infection history

**DOI:** 10.1101/2024.04.07.24305311

**Authors:** Nyall Jamieson, Christiana Charalambous, David M. Schultz, Ian Hall

## Abstract

Understanding the temporal relationship between key events in an individual’s infection history is crucial for disease control. Delay data between events, such as infection and symptom onset times, is doubly censored because the exact time at which these key events occur is generally unknown. Current mathematical models for delay distributions rely solely on heuristic justifications for their applicability. Here, we derive a new model for delay distributions, specifically for incubation periods, motivated by bacterial-growth dynamics that lead to the Burr family of distributions being a valid modelling choice. We also incorporate methods within these models to account for the doubly censored data. Our approach provides biological justification in the derivation of our delay distribution model, the results of fitting to data highlighting the superiority of the Burr model compared to currently used models in the literature. Our results indicate that the derived Burr distribution is 13 times more likely to be a better-performing model to incubation-period data than currently used methods. Further, we show that incorporating methods for handling the censoring issue results in the mean of the underlying continuous incubation-period model being reduced by a whole day, compared to the mean obtained under alternative modelling techniques in the literature.

**Author summary:** In public health, it is important to know key temporal properties of diseases (such as how long someone is ill for or infectious for). Mathematical characterisation of properties requires information about patients’ infection histories, such as the number of days between infection and symptom onset, for example. These methods provide useful insights, such as how their infectiousness varies over time since they were infected. However, two key issues arise with these approaches. First, these methods do not have strong arguments for the validity of their usage. Second, the data typically used is provided as a rounded number of days between key events, as opposed to the exact period of time. We address both these issues by developing a new mathematical model to describe the important properties of the infection process of various diseases based on strong biological justification, and further incorporating methods within the mathematical model which consider infection and symptom onset to occur at any point within an interval, as opposed to an exact time. Our approach provides more preferable results, based on AIC, than existing approaches, enhancing the understanding of properties of diseases such as Legionnaires’ disease.

## Introduction

In epidemiology, the temporal relationship between key events in an individual’s infection history is important to understand. For example, a disease that has a long delay from infection to onset of infectiousness may be amenable to contact tracing, and the relationship between these two events can be important for disease control [1, 2]. Often these events are a simplification of a continuous process (i.e., infectivity may not start or end at specific times but instead increase and then decrease over time). For diseases such as Legionnaires’ disease, which spread via airborne dispersion from environmental sources (rather than person-to-person contact), characterisation of the incubation period is critical for source identification (or reverse epidemiology).

Here, we consider the time from infection to symptom onset. The relationship between viral or bacterial load in one’s body and onset of symptoms can be difficult to describe. In brief, the presence of a virus or bacteria within an individual results in an inflammatory immune response that leads to an observable response of symptoms. An exact mathematical model accurately describing the infection process is not feasible to develop due to the large number of different cytokines and cell interactions involved in the immune response, as well as a lack of a clear understanding of how the pro-inflammatory cytokines relate to the appearance of symptoms and a lack of data to parameterise each specific process in the immune response. Previous models for the incubation period provide parsimonious simplifications of the infection process, and include in-host models (often assuming symptom onset is proportional to bacterial load [3]), through to simpler probability models (justified on model parsimony or computational capacity). In the latter case, popular distributions include the gamma, log-normal and Weibull distributions [4–6].

The validity of these distributions has not been explored, and application is based solely on heuristic justification. The arguments for common distributions can be described as follows. The gamma distribution is the sum of *n* exponentially distributed random events, and so fitting to data can help inform the structure of compartmental models [7]. The log-normal distribution is a skewed distribution often applied to biological processes in which the process mean time is relatively low, but its variance is large and results from taking the exponential of a series of normally distributed events. Finally, the Weibull distribution is a classic reliability-theory distribution where the hazard of an event occurring is strictly monotonic over time.

To illustrate the heuristic justification of distributions, we consider Legionnaires’ disease and the statistical analysis that has been conducted in the literature for studying the incubation period. In this case, several papers have used a range of days (2–10) prior to symptom onset and consider all days in this period as a potential infection date [8–14]. Alternatively, others have assumed a median incubation period of either five days [15] or seven days [16], with infection dates obtained by subtracting the median from the symptom onset date. Another common approach is to consider a gamma-distributed incubation period [17]. All papers that take this approach have followed the ideas and method proposed in [4] using a gamma distribution to describe an outbreak in Melbourne [18].

One issue arising is that incubation-period data is given as an integer number of days, implying that each case becomes infected at the same moment from the exposure, and that symptoms develop in an integer amount of days. To illustrate this issue, take two cases in which symptom onset occurs the day after infection. The individual could have been infected at 11:59pm and became symptomatic at 00:01am the next day, or alternatively they could have been infected at 00:01am and became symptomatic at 11:59pm the next day. These two scenarios are 2 minutes and 1 day, 23 hours, 58 minutes long, respectively, but they both correspond to one integer day in the dataset. These simplifications give a lower resolution of the time delay between these events due to lack of knowledge of the exact infection and symptom onset times. Essentially, continuous distributions are being fitted to discretized versions of continuous data, and the result is interval data with censored start and end times.

This type of discretized data is commonly used for analysis without consideration for the censoring issue. Using standard probability distributions, as well as censored incubation-period data in statistical analysis, is likely to produce biased inference. Using incubation-period data expressed as an integer number of days will likely lead to a false understanding of delays between key events for specific diseases, such as the incubation period, and produce incorrect conclusions. A model describing the incubation period of Legionnaires’ disease has been built with this type of data [4], but the model is flawed and can be improved upon by accounting for the issues mentioned above. There are various ways to handle the censoring issue, which we discuss in the next section.

In this paper, a new model for incubation periods is derived with potentially stronger justification for its validity than methods currently used in the literature. We apply our new model to a variety of diseases to provide statistically significant improvements compared to currently accepted and used models. We also apply techniques that remove the bias from fitting models to censored data and allow for reliable model-fitting, providing a new understanding of the incubation periods of various diseases. We apply these methods to anthrax, salmonellosis and campylobacteriosis, as well as taking a specific focus on Legionnaires’ disease to illustrate the typical kind of improvement achievable with these methods. For the successful models, we develop some distribution theory, calculating their moments and quantile functions, which can be found in S1 Appendix in the Supplementary Material.

## Materials and methods

In this section, we develop methods for handling both of the problems discussed in the introduction. First, we adapt the methods developed in [19] for use on incubation-period data in order to account for its censored nature. Second, we consider a probabilistic approach to develop a new model for incubation periods of diseases. We assume exponential growth of bacteria early after infection, as well as a further assumption of the probability of symptom onset being proportional to the bacterial load within an individual until saturating once some load has been reached. Third, we discuss the methods for analysing our fitted models and how we determine which model performs better, so that we can conclude whether or not our developed model offers more reliable results than using methods currently developed in the literature. Finally, we introduce the data used for incubation-period analysis and discuss the reasons why this data is considered censored.

### Doubly interval-censored modelling

Methods for handling censored data in epidemiological studies have been proposed in the literature to develop discrete analogues of continuous distributions that preserve properties of their continuous counterparts [20]. However, most of these methods are either too simple, do not result in valid probability mass functions, or assume that infection occurs exactly at midnight.

The exact time at which symptoms occur in an individual can not be determined based on when they reported their illness to authorities. Similarly, the exact time at which an individual becomes infected is also difficult to ascertain. We need a method for handling the fact that these times are unknown (i.e., to account for the uncertainty within a model), so that analysis of any subsequent models is reliable. To consider doubly censored data, a natural approach is to forget the assumption that the exact infection and symptom onset times are known and introduce a time period in which these two events may occur, with a probability distribution for the occurrence within this period [19]. The method proposed in [19], which considers doubly interval-censored (DI) data, is described as follows.

Define *T* and *S* to be the time of infection and symptom onset respectively (with *t* and *s* being realisations of these random variables respectively), and *Z* = *S* – *T* as the incubation period of the infection. Consider two intervals where *T* and *S* could lie within because the exact times of *T* and *S* are not known. In other words, let *T* ∈ (*T*_*L*_, *T*_*R*_) and *S* ∈ (*S*_*L*_, *S*_*R*_). The incubation period *Z* is given as a random variable with p.d.f. *f* (*s* – *t*) (Fig. 1).

**Fig 1.**
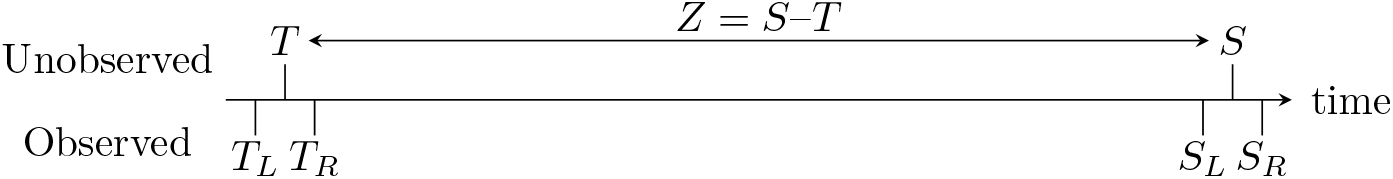
Diagram visualising the doubly interval-censoring method [19], highlighting the data typically observed, but accounting for the fact that infection and symptom onset times are not observed exactly and intervals of possible times must be considered.

The p.d.f. of *T* is defined as *f*_*T*_ (*t*) and the p.d.f. of *S* is defined as *f*_*S*_(*s*). The time at which a person becomes infected and the time taken from infection to symptom onset are independent, which leads to *f*_*S*_(*s* |*t*) = *f* (*s* – *t* | *t*) = *f* (*s* – *t*). Finally, define the joint p.d.f. of *T* and *S* as

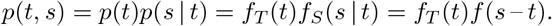

From this, the likelihood for a doubly interval-censored observation *x* is derived.

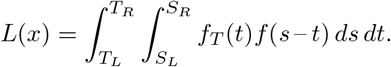

To implement methods found in [19] to incubation-period data, the following approach is taken. Because the data is rounded to the nearest day, a natural assumption is that *T*_*L*_ = 0 and *T*_*R*_ = 1, so infection occurs at any point on the infection date. Defining *x* to be the number of days from exposure to symptom onset, set *S*_*R*_ = *x* and *S*_*L*_ = *x* – 1, so that the symptoms develop at some point on the stated date of symptom onset. There is not much evidence to indicate what distribution *f*_*T*_ (*t*) might be, so a reasonable assumption would be to let *f*_*T*_ (*t*) be uniform (i.e., *f*_*T*_ (*t*) = 1 on *t* ∈ (0, 1), 0 otherwise). Other options could be to permit a lower chance during nighttime or a higher chance when people are outdoors, but these will depend on specific release scenarios and are not likely particularly identifiable in data. As *f* (*s* – *t*) is the p.d.f. of the incubation period, the log-likelihood is calculated as follows:

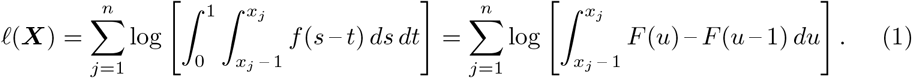

In the next section, we develop various distributions to describe the incubation period, and later fit the doubly interval-censored model to these distributions, to determine which one provides the most optimal fit.

### Derivation of the incubation period model

Incubation period data describes the cases who become symptomatic. Given the knowledge that all individuals in the data will become symptomatic, this section discusses different mathematical models for the occurrence of symptoms onset within a population. We explore how the results for these different methods link and we develop a new model for incubation periods, by starting from a probabilistic approach of symptom onset occurrence.

#### A probability-based approach

A mathematical model can be built considering probabilities of symptom onset occurrence. Define *N*(*t*) as the population of individuals who are infected, but are not yet symptomatic at time *t*, and *Q*(*t*) as the population of individuals who are symptomatic at time *t* with *N*(0) = *N*_0_ and *Q*(0) = 0 and *Q*(*t*) + *N*(*t*) = *N*_0_, ∀*t* ∈ ℝ^+^. Next, assume that there is a probability *p*(*t*) that a not-yet-symptomatic individual will start to experience symptoms at a point in time *t*, then 1 – *p*(*t*) will be the probability that the individual will remain asymptomatic. Hence (1 – *p*(*t*))^*N*(*t*)^ is the probability that nobody who is not-yet-symptomatic will start experiencing symptoms at this point in time, and (1 – *p*(*t*))^*N*(*t*)*δt*^ is the probability that nobody new will experience symptoms in a time increment *δt*. Following this, define *δQ*(*t*) = 1 – (1 – *p*(*t*))^*N*(*t*)*δt*^ to be the probability that there is at least one individual who starts to experience new symptoms in time increment *δt*. By writing *µ*(*t*) = – log(1 – *p*(*t*)), the probability of any new symptom onset appearance can be written as *δQ*(*t*) = 1 – *e*^−*µ*(*t*)*N*(*t*)*δt*^. Using a Taylor expansion on the exponential term, dividing by *δt*, and taking the limit *δt* → 0 changes this probability to a rate as follows:

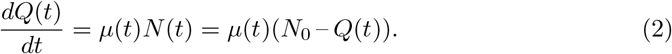

This approach leads to a separable ordinary differential equation analogous to the cumulative distribution of the exponential distribution with a time-varying rate parameter.

It can be deduced that 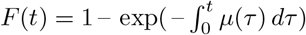 and that 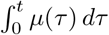 is the accumulated hazard. Hence the rate of symptom onset, *µ*(*t*), is the hazard function of an individual becoming symptomatic. Therefore, the hazard of an individual becoming symptomatic at a point in time is equal to the rate of symptom onset at that time. The scenario discussed here can be considered from an inhomogeneous Poisson-process perspective, and the results of the hazard are identical to the inhomogeneous exponentially distributed model. It can be noted here that if *µ*(*t*) is constant that this would lead to the exponential distribution and if *µ*(*t*) ∝ *t*^*a*^ for some constant *a* this would suggest the incubation period is a Weibull distributed random variable. The gamma distribution arises by assuming the incubation period is the sum of a number of stages of constant length *µ*.

However, symptom onset is likely proportional to bacterial load at low loads (i.e., the early stages of infection) before saturating at large loads. The bacterial population early after infection will be approximately some exponential function of time [3, 21, 22]. Therefore, the left tail of the c.d.f. of the incubation-period distribution is given by some function 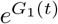, whilst in the later stage, the c.d.f. should tend to 1 exponentially given by a function *G*_2_(*t*), as is the case of the hazard function above. Mathematically, with a median *T*, and considering the case where *G*(*t*) = *G*_1_(*t*) = *G*_2_(*t*), an equation for the c.d.f. that satisfies these conditions is given as follows:

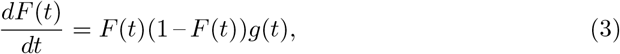

where 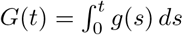 *ds* for some function *g*(*s*). The ODE that arises in (3) defines the Burr family of distributions and is discussed in further detail in the next section.

#### Burr distribution

A *Burr* distribution is a distribution whose c.d.f., *F*(*t*),

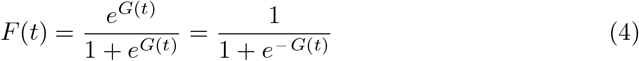

is the solution of (3). Theoretically, there are no constraints on *G*(*t*) in (4). Twelve main distributions within the Burr family have been characterized [23], named as Burr type I, Burr type II, up-to Burr type XII, but we only consider Burr distributions defined over a domain of (0, ∞).

Some delay distributions arising in epidemiology do permit negative values. For example, the time from symptom onset in infector to symptom onset in infectee could be negative. In this paper we limit consideration to strictly positive cases. A negative incubation period is not possible, nor is a fixed upper limit constraint expected. The only biologically feasible distributions are types III, X and XII. The type III distribution could be derived from the flexible generalized gamma distribution with the scale parameter following an inverse Weibull distribution [24]. Similarly, type XII could be derived from the Weibull distribution where the scale parameter follows an inverse generalized gamma distribution [24].

The Burr distributions and the gamma distribution have parameters which share the same symbols for notational simplicity, although they have different interpretations and their fitted estimates can not be directly compared. To avoid confusion, we provide a subscript for each parameter to clarify which distribution this parameter corresponds to (i.e., *α*_*III*_ for the *α* parameter in the type III Burr model) in the text but drop this in tables and figures for brevity. Further, we note that the types III, X and XII used in this research are a generalization of types III, X and XII Burr distributions used in the literature [23], where the time variable is scaled by an additional parameter. Type X is defined with two variables that provide models as parsimonious as the three previously trialled: gamma, log-normal, and Weibull. Further, both type III and XII have a scale parameter *γ*_*III,XII*_ and two shape parameters *α*_*III,XII*_ and *β*_*III,XII*_.

#### General derived Burr distribution

In (3) *g*(*t*) has a physical interpretation; the function tends to the rate of symptom onset, *µ*(*t*), in individuals at a time *t* as *t* increases. Given *F*(*t*) = (1 + *e* ^− *G*(*t*)^)^*−*1^, in general, then *G*(*t*) → *t/β*_*D*_ (or *g*(*t*) → 1*/β*_*D*_) for some constant *β*_*D*_ as *t* → ∞ on the basis that relatively long incubation periods are memory-less Markovian random variables. In principle, *F*(0) = 0, so *G*(*t*) → –∞ for *t* → 0 (or *G*(0) is very large if not actually infinite). Taking the above into account, we propose *g*(*t*) = 1*/β*_*D*_ + *α*_*D*_*/t*, and as such *G*(*t*) = *t/β*_*D*_ + *α*_*D*_ log(*t*) + *C*, where *C* is a constant of integration. We define *T*_*D*_ as the median, which satisfies *G*(*T*_*D*_) = 0. Hence, *C* = – *T*_*D*_*/β*_*D*_ – *α*_*D*_ log(*T*_*D*_) and thus

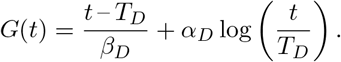

Equations for the c.d.f. and p.d.f. for the derived Burr, as well as the gamma and other Burr distributions, are given in Table 1. As discussed, *T*_*D*_ is the median of the distribution. The reciprocal of *β*_*D*_ is the eventual Markovian rate of symptom onset in individuals for *t* ≫ *T*_*D*_. Additionally, there are two details worth noting when analysing the physical interpretation of *α*_*D*_. First, *α*_*D*_ is an exponent of *t* controlling the growth of probability, as 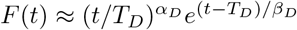 for *t* ≪ *T*_*D*_. Second, the general derived Burr distribution approaches the exponential distribution for *t* ≫ *T*_*D*_. The rate at which the derived Burr approaches the exponential distribution increases for decreasing *α*_*D*_. Therefore, the parameter *α*_*D*_ can be interpreted as a parameter that limits the rate at which the symptom onset process in an individual becomes Markovian. Finally, all parameters must be strictly greater than zero.

**Table 1.**
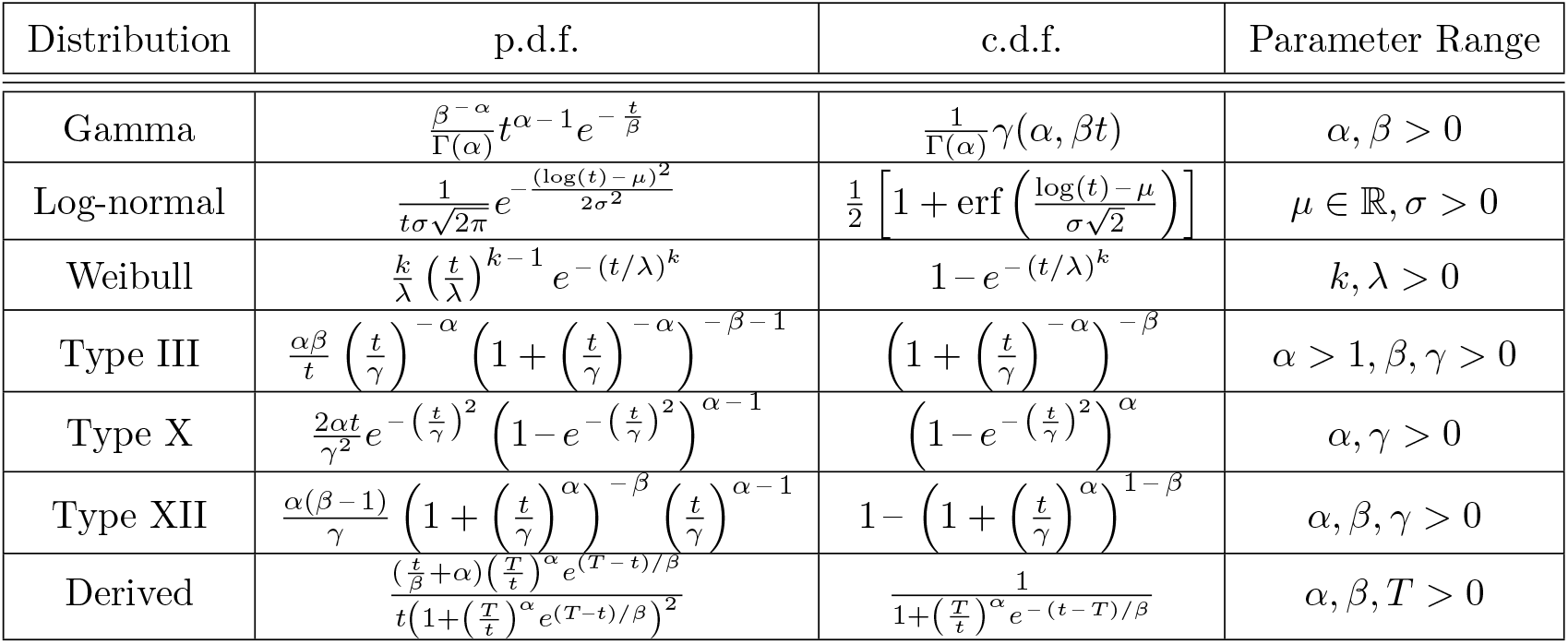
The Burr distributions valid over (0, ∞) and previously trialled distributions [4] with their corresponding p.d.f and c.d.f., as well as the parameters in each model.

### Model comparison

We fit each type of Burr distribution to the data, and assess all the models in terms of their goodness of fit in comparison to the more widely used gamma distribution. There are various criteria that penalise models to varying degrees and judge models from different perspectives, such as from an information theory view-point or an expected loss view in decision theory. The most commonly used methods for model selection are the Akaike information criterion (AIC) and Bayesian information criterion (BIC). The AIC and BIC share a similarity in that the aim of a good model is to minimize their score. Generally, AIC puts more emphasis on good model prediction, whereas BIC favours model parsimony [25]. Because our goal is good model prediction, the AIC will be used in deciding desirable model fits.

By defining *p* as the number of parameters in the model and *ℓ*(***X***) as the log likelihood of that model given the data ***X***, the method to calculate the AIC is given as follows:

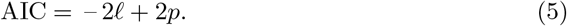

Additionally, we consider the difference between AIC values and the minimum AIC, [26] defined this difference as Δ_*i*_(AIC) = AIC_*i*_ – min(AIC), which is then used to calculate the Akaike weights [26]:

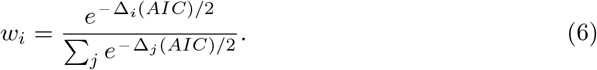

When fitting models to data to compare the validity of a Burr distributed model over the gamma distributed model, the weights *w*_*i*_ can be interpreted as the probability that model *i* is the best model, given the data and set of models being considered [26]. Furthermore, the ratio *w*_*i*_*/w*_*G*_, where *w*_*i*_ is the weight for the *i*^th^ model and *w*_*G*_ is the weight of the gamma distributed model, can be interpreted as how much more likely model *i* is the best fitting model compared to the gamma model. Alternatively, we also derive the normalized probability that the *i*^th^ model is preferable to the gamma model, given by *w*_*i*_*/*(*w*_*i*_ + *w*_*G*_).

The final method of comparison considered is the Bayes factor. The maximum likelihood estimates that we obtain in analysis can be considered maximum a posteriori estimates with a uniform prior and are used in this context for conducting the Bayes factor calculations. The ratio of likelihoods of two models determines whether there is enough evidence to prefer one model to another. Let *ℓ*_*A*_ and *ℓ*_*B*_ be the likelihood of two models, *A* and *B* respectively. We calculate the value

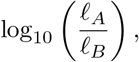

which is used for comparison between models. Based on [27], if this value is in the range (0, 0.5), there is little evidence that model A outperforms model B, (0.5, 1) gives substantial evidence that model A outperforms model B, (1, 1.5) gives strong evidence that model A outperforms model B and the larger the value, the stronger the evidence that model A outperforms model B. This method shall be used to compare each of the Burr distributions separately with the gamma model.

### Incubation-period data

To test these models, we employ incubation-period data from an outbreak of Legionnaires’ disease in Melbourne in April 2000 [18]. The data for the Melbourne outbreak contains the number of days taken for each Legionnaires’ disease case to develop symptoms from their exposure date, and several potential distributions for fitting the data have been compared [4]. The results indicated that the gamma distribution provided the best fit [4] out of their proposed models.

Further, we gather incubation-period data for anthrax, campylobacteriosis and salmonellosis for analysis. The anthrax outbreak in 1979 contains data for the known incubation-periods of patients [28]. A literature review has been conducted analysing different salmonellosis studies that contain full data of the incubation periods [29]. Awofisayo-Okuyelu et al. [29] noticed that the incubation periods varied between studies. They grouped studies into subsets using a clustering process, in which the grouped studies did not have any statistically significant difference in their incubation-period data. Similarly, Awofisayo-Okuyelu et al. [30] conducted a review for campylobacteriosis in which the incubation periods varied between studies, and they combined datasets which were not statistically significantly different using a clustering process similar to [29]. We provide an Excel sheet of the incubation-period data for these other diseases in S4 Data in the Supplementary Material.

The data gathered for these diseases share a similarity with the Legionnaires’ disease data, in that the data contains the integer number of days taken for each case to develop symptoms. The fact the data for all of these diseases contains integer days implies that each case takes an exact multiple of 24 hours from infection to the appearance of symptoms, which is not realistic. If we assume that the dates of infection and symptom onset are accurate, then we know the date of these events, but the specific times on the given days are unknown. We are dealing with doubly censored data.

## Results

Now that we have developed the Burr distribution as an incubation-period model based upon biological justifications, the next step is to fit these models to the incubation-period data of various diseases. We begin by fitting the incubation-period models to the Legionnaires’ disease data, to draw comparisons between the models’ performance. Next, we conduct the same analysis on other diseases such as anthrax, campylobacteriosis and salmonellosis. Finally, we conduct a simulation in which incubation-period data is fabricated. We compare the results from fitting the incubation-period models to this data, as we compare the parameter estimates obtained from fitting the gamma and derived Burr distributions to this data in an attempt to assess the relationship between these parameters.

### Analysis of the Melbourne data

The gamma distribution is currently most frequently used to model Legionnaires’ disease incubation periods [4], thus we produce models using a gamma distributed incubation period, as well as a Burr distributed incubation period, to allow for comparison between the two. Models are fitted using both the continuous and doubly interval-censored models to offer comparison between the two methods.

This section begins by providing the results from fitting the incubation-period models to the data (Table 2). Comparisons are drawn both between the incubation-period models, as well as between model-fitting approaches and the effect that has on our understanding of Legionnaires’ disease incubation periods. We provide analysis of the moments of these Legionnaires’ disease incubation-period models in S1 Appendix in the Supplementary Material. Further, in this appendix we provide visual comparison of the accumulated hazard of these models for large time, to examine their ability to accurately display a Markovian property of long incubation periods. The analysis and production of plots was conducted on R, with the code provided in S3 Code in the Supplementary Material.

**Table 2.** Results from fitting the gamma and four Burr distribution models to the Melbourne incubation-period data using both the continuous and DI likelihood fitting methods.

| Method | Analysis | Distribution |  |  |  |  |
| --- | --- | --- | --- | --- | --- | --- |
| - | - | Gamma | Burr III | Burr X | Burr XII | Derived |
| Continuous | Parameter estimates (s.e) | $\alpha = 4.963$<br>(0.636)<br>$\beta = 1.275$<br>(0.171) | $\alpha = 5.664$<br>(0.970)<br>$\beta = 0.444$<br>(0.126)<br>$\gamma = 7.690$<br>(0.642) | $\alpha = 1.525$<br>(0.203)<br>$\gamma = 6.054$<br>(0.335) | $\alpha = 2.955$<br>(0.366)<br>$\beta = 4.451$<br>(2.309)<br>$\gamma = 10.031$<br>(3.131) | $\alpha = 1.725$<br>(0.751)<br>$\beta = 2.738$<br>(0.989)<br>$T = 6.050$<br>(0.254) |
|  | ML | -272.66 | -270.92 | -271.81 | -271.15 | -270.84 |
|  | AIC | 549.32 | 547.83 | 547.62 | 548.29 | 547.67 |
| | $\omega/\omega_G$ | 1 | 2.106 | 2.340 | 1.674 | 2.282 |
| | $\omega/(\omega + \omega_G)$ | - | 0.678 | 0.701 | 0.626 | 0.695 |
|  | BF | - | 0.756 | 0.369 | 0.656 | 0.790 |
| DI | Parameter estimates (s.e) | $\alpha = 3.479$<br>(0.228)<br>$\beta = 0.653$<br>(0.009) | $\alpha = 5.475$<br>(1.048)<br>$\beta = 0.334$<br>(0.009)<br>$\gamma = 7.229$<br>(0.415) | $\alpha = 1.065$<br>(0.020)<br>$\gamma = 5.848$<br>(0.134) | $\alpha = 2.249$<br>(0.083)<br>$\beta = 8.643$<br>(98.094)<br>$\gamma = 14.194$<br>(105.311) | $\alpha = 0.880$<br>(0.320)<br>$\beta = 2.110$<br>(0.315)<br>$T = 5.075$<br>(0.067) |
|  | ML | -274.33 | -271.42 | -272.59 | -272.27 | -270.75 |
|  | AIC | 552.66 | 548.84 | 549.18 | 550.54 | 547.50 |
| | $\omega/\omega_G$ | 1 | 6.753 | 5.697 | 2.886 | 13.197 |
| | $\omega/(\omega + \omega_G)$ | - | 0.871 | 0.851 | 0.743 | 0.930 |
|  | BF | - | 1.264 | 0.756 | 0.895 | 1.555 |

When fitting using the continuous maximum likelihood method, types III, X, XII and the derived Burr perform better than the gamma regardless of which scoring criterion is used. Because type X is a two-parameter distribution, the fact that its maximized log-likelihood is higher than gamma’s automatically means that its minimized AIC will be lower. Types III, XII and the derived Burr perform better than gamma depending on how harshly they are penalized for their extra parameter. Based on AIC, our ideal information criterion for model selection, these perform better than the gamma distribution. On the whole, all Burr distributions perform better than the gamma distribution. From considering the Akaike weights ratio *w/w*_*G*_, the derived Burr, type III, and type X are at least two times as likely to be a better-performing model than the gamma distributed model. Additionally, each Burr model provides at least a 62% chance of being a better fitting model than the gamma distributed model, with the derived Burr model being 70% more likely to be better than the gamma model. Looking at the Bayes factor, there is no substantial evidence to favour type X over the gamma. However, this criterion gives substantial evidence that both types III, XII as well as the derived Burr are all favourable over the gamma distribution.

Next, when fitting using doubly interval-censoring methods, type X again outperforms the gamma distribution. Types III, XII and the derived Burr perform better than the gamma model, based on AIC, even with one extra parameter. When considering the Akaike weights, all the Burr distributed models perform much better than the gamma distribution, with the derived Burr being over 13 times more likely to be the better-fitting model. Additionally, when considering *w/*(*w* + *w*_*G*_), all Burr models are more likely to be perform better than the gamma distribution, with the derived Burr being 93% likely. Finally, the Bayes factor for types X and XII both show substantial evidence of a better fit than the gamma distribution. Further, the Bayes factor for type III and the derived Burr both show strong evidence of a better fit than the gamma model.

The same conclusions are drawn regardless of maximum likelihood fitting method; all the distributions provide a better fit than the gamma distribution. Results from using the DI methods agrees with the continuous likelihood method in that *β*_*XII*_ and *γ*_*XII*_ in the Burr type XII model have large standard errors, indicating that they are not important in the model fitting procedure.

When fitted using continuous maximum likelihood methods, all Burr distributions considered offer a similar curve when plotted, as expected, but do vary slightly as to the model value or the value of the p.d.f. at the mode (Fig. 2). The Weibull distribution provides a similar modal value for the incubation period, but is more variable than the Burr models. The gamma distribution provides a slightly lower modal value than the Burr models. The log-normal model provides a noticeably different curve to the Burr models and provides a much lower modal incubation period, with a lighter left tail and heavier right tail than all the other distributions.

**Fig 2.**
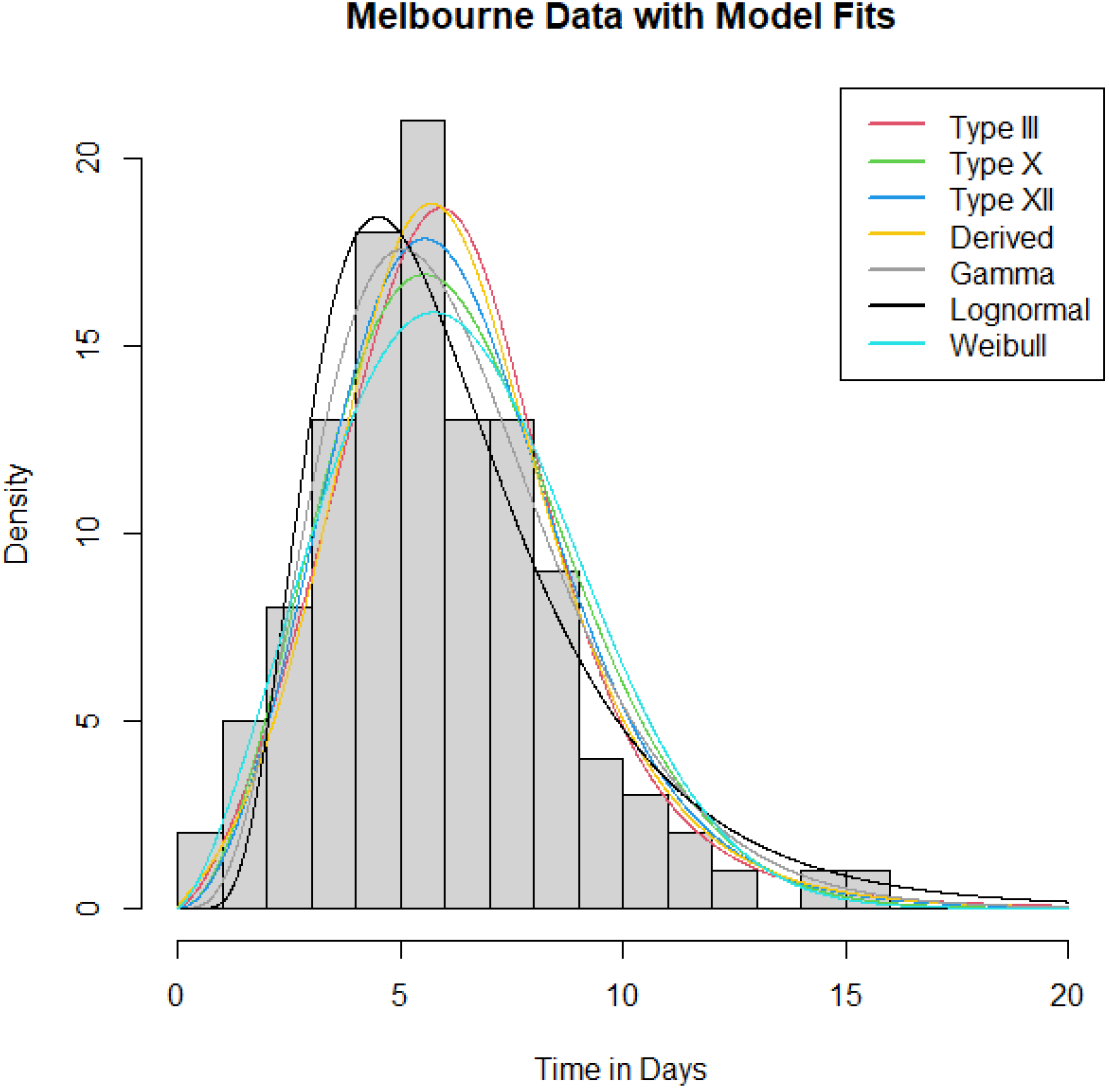
A plot of the Melbourne case data with the four fitted Burr distributions included, which offer a visual representation of the incubation-period distributions trialled.

The mean of each fitted distribution along with a bootstrapped 95% confidence interval is calculated under both the continuous method and the doubly interval-censored method to identify any differences across distributions and across methods, and is provided in Table 1 of S2 Figure in the Supplementary Material. A common theme exists, which is that, for each distribution, the mean for the doubly interval-censored model is approximately a day less than the continuous model (5.3 days compared to 6.3 days), with the confidence intervals for each model having no overlap across all the distributions. These results are statistically significant and provide support for using a doubly interval-censored model to more accurately represent the incubation period of Legionnaires’ disease.

For all of the distributions, the density under the doubly interval-censored approach is shifted more towards the left, indicating that the incubation period is shorter than when just taking the incubation period as exact integer days (Fig. 3). Indeed, the doubly interval-censored methods account for a potential delay between exposure time and infection as well as a delay between symptoms starting to develop and the person reporting the symptoms, whereas the continuous model does not account for either delay, resulting in longer times for the incubation periods.

**Fig 3.**
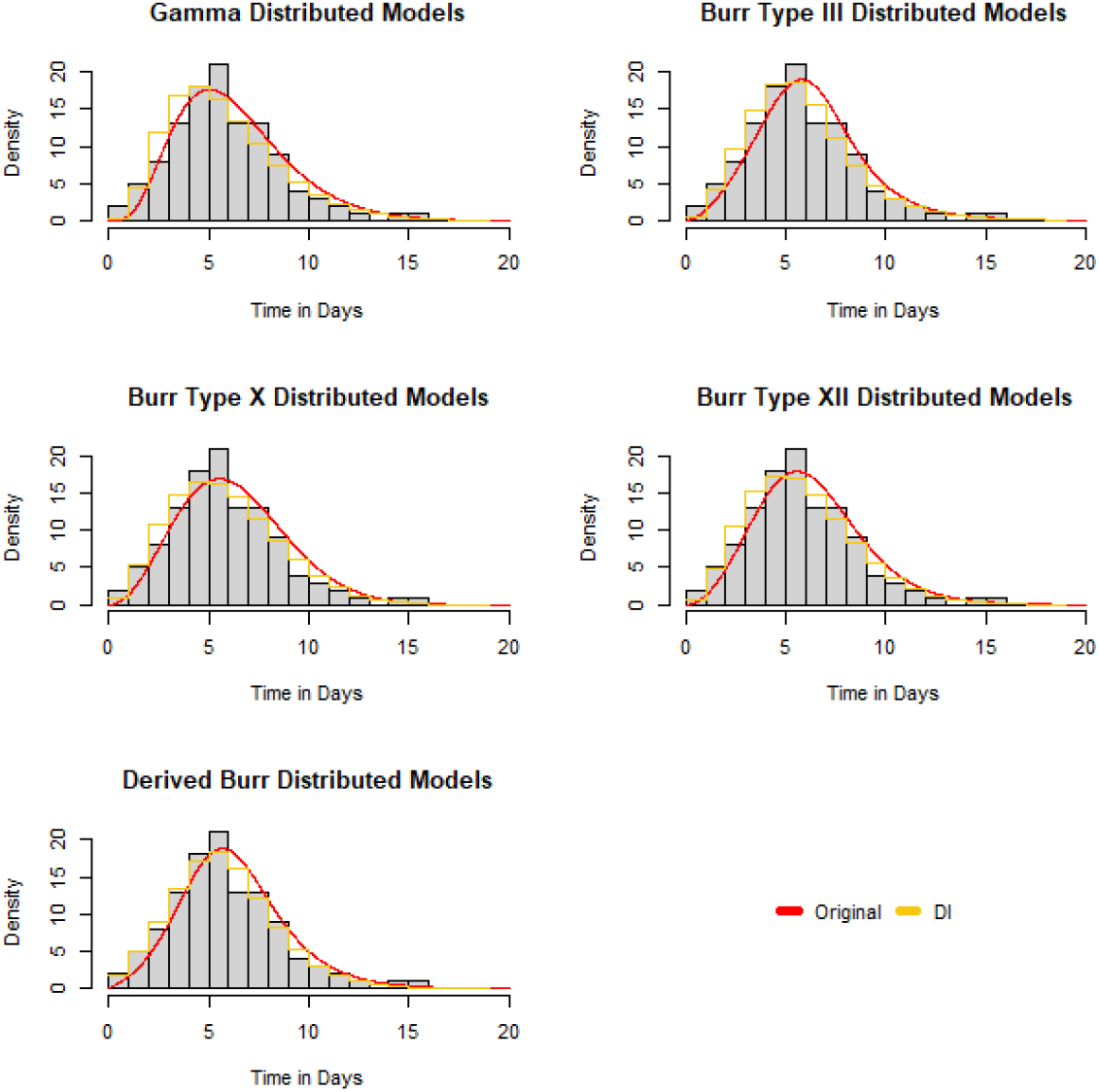
Plots of the Melbourne data with the continuous model fits in red and the doubly interval-censored model fit as a step function in yellow. Each step of the function is a horizontal line from *t* ∈ (*a, b*] where *a* = ⌊*t*⌋ and *b* = ⌈*t*⌉.

### Application to other diseases

To further check the validity of the Burr distribution, we fit the doubly interval-censored models to data of the incubation periods for different diseases: anthrax [28], campylobacteriosis [30] and salmonellosis [29]. Figures of resulting model fits provided in S2 Figure in the Supplementary Material, along with the obtained parameter estimates and standard errors of these estimates contained in Table 1 of S2 Figure in the Supplementary Material. We use both the continuous and the doubly interval-censored methods to fit the gamma and the Burr distributions, to compare which model provides a better fit (Table 3).

**Table 3.** Comparing Burr and gamma models on anthrax, salmonellosis and campylobacteriosis datasets. For this table: Y represents that the given model outperforms the gamma distribution, N represents that the given model does not outperform the gamma distribution and H represents that the given model outperforms the gamma distribution based on maximum likelihood, but not on AIC.

| Disease | Method | Distribution |  |  |  |
| --- | --- | --- | --- | --- | --- |
|  |  | Burr III | Burr X | Burr XII | Derived |
| Anthrax | DI | Y | N | H | H |
|  | Continuous | Y | N | Y | H |
| Salmonellosis | DI | HYYY | YYNN | YYYY | HYYY |
|  | Continuous | NYYN | YYNN | YYYY | HYYY |
| Campylobacteriosis | DI | YNNYY | NNYNY | HNNYY | YHNYY |
|  | Continuous | YYNYY | NNNNY | YYNYY | YYNYH |

Burr types III and X offer mixed results across datasets and do not consistently outperform the gamma distribution. Apart from the third campylobacteriosis dataset, both the derived Burr and type XII Burr models consistently outperform the gamma distribution. When comparing optimal fits across datasets, the derived Burr appears to be the most optimal out of these choices of models.

We note that there is no clear pattern between any of the fitted *α*_*D*_ and *β*_*D*_ parameter estimates and the performance of the derived Burr distribution. Additionally, there is no clear pattern from the anthrax, campylobacteriosis and salmonellosis datasets as to whether the estimate of the median, *T*_*D*_, relates to the performance of the derived Burr model. However, the lack of sensitivity for *T*_*D*_ is logical as *T*_*D*_ solely scales the distribution about the median, and the ability of the derived Burr distribution to fit well to incubation period data will depend more on the tails in the curve and around the median, as opposed to the median itself.

We can draw conclusions on which scenarios the derived Burr distribution will outperform the gamma distribution based on plots provided in Figure 1 of S2 Figure of the Supplementary Material. The third campylobacteriosis dataset was the only dataset in which the derived Burr did not outperform the gamma distribution based on either maximized likelihood or on AIC. This dataset is unique in that the incubation period ranges from one to five days. As a result, the effect of the censoring bias will be much larger, due to the fact that this incubation period is much shorter. Therefore, this is not an ideal dataset to use to assess model performance.

Next, we consider the datasets in which the derived Burr outperformed the gamma distribution based on maximized likelihood but not on AIC, regardless of model fitting procedure. The anthrax dataset has a high density after the mode and does not tail off, and the probability distribution of the first salmonellosis dataset does not have a clearly defined mode and is negatively skewed. The derived Burr distribution offers close results to the gamma distribution when it comes to modelling incubation periods without a clear mode or tail off in probability of illness, but is a better-performing distribution when this structure is clearer defined.

Finally, fitting to the second and fifth campylobacteriosis datasets resulted in the derived Burr outperforming the gamma on maximized likelihood but not on AIC. The incubation period for these datasets is relatively small, meaning that the bias from the censoring issue is large when fitting models to these datasets. The campylobacteriosis datasets that resulted in the derived Burr distribution outperforming the gamma distribution were the ones in which the modal time was clearly defined and not a wide range of times at the peak of the distribution. This further supports the hypothesis that the derived Burr becomes more preferable when either the mode is more apparent, or the range of incubation periods in the datasets is not too short that the censoring becomes a larger issue.

### Results of model-fitting to simulated data

We now further assess the validity of the Burr distributions by comparing their fits, along with those of the gamma distribution, to fabricated data. Specifically, we aim to analyse how the parameter estimates of the gamma distribution relate to the parameter estimates of the derived Burr distribution for different datasets, to gain a further understanding of how the derived Burr parameters can be interpreted.

Initially, we generate a sample of size 1000 from a gamma distribution with given shape *α*_Γ_ and mean *µ*_Γ_ (scale *β*_Γ_ = *µ*_Γ_*/α*_Γ_). Then the derived Burr parameter estimates are obtained from fitting to this dataset by continuous maximum likelihood, so that analysis can be conducted on the effect that varying *α*_Γ_ ∈ (0, 5) or *µ*_Γ_ ∈ (1, 20) has on these estimates. A heatmap is produced to visualise this effect (Fig. 4).

**Fig 4.**
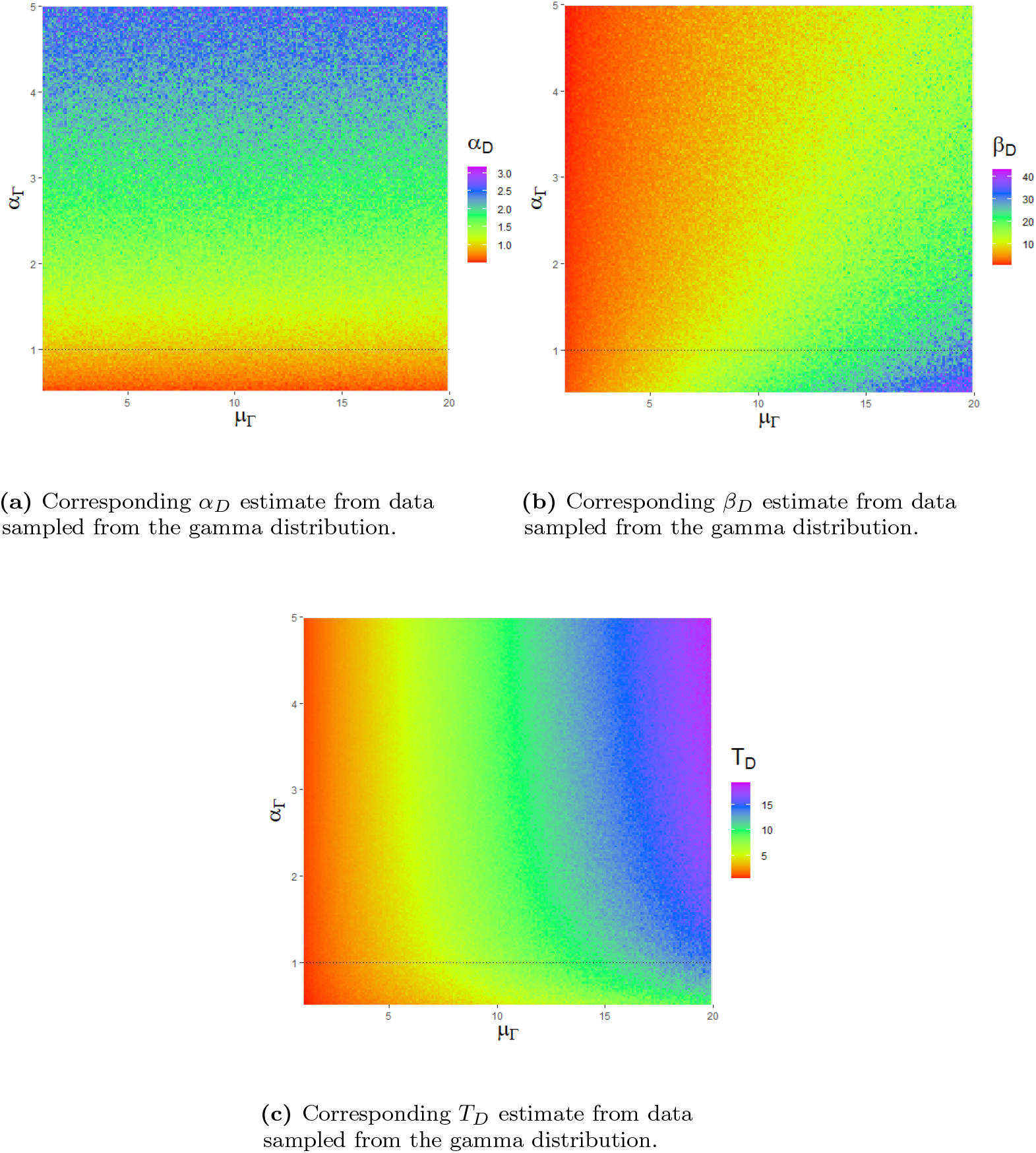
Heatmap of the results from the third simulation. The derived Burr distribution is fitted to data generated from the gamma distribution with parameters *α*_Γ_ and *µ*_Γ_, with the obtained derived Burr parameter estimates plotted.

Parallels exist between the interpretations of *α*_Γ_ and *α*_*D*_. Increasing *α*_Γ_ results in a larger discrepancy between the gamma distribution and the exponential distribution. Thus, *α*_Γ_ limits quickly the distribution becomes Markovian over time. Therefore, a positive correlation between *α*_Γ_ and *α*_*D*_ is expected (Fig. 4a). The results indicate that *µ*_Γ_ does not have an effect on the rate at which the gamma distribution becomes Markovian.

Similarly, parallels exist between the interpretations of *β*_Γ_ and *β*_*D*_. The hazard rate for the gamma distribution tends to 1*/β*_Γ_ as *t* → ∞. Hence, 1*/β*_Γ_ as the eventual Markovian rate of symptom onset for this distribution. Thus, a positive correlation between *β*_Γ_ and *β*_*D*_ is logical (Fig. 4b). Therefore, the effect that varying either *µ*_Γ_ or *α*_Γ_ in *µ*_Γ_ = *α*_Γ_*β*_Γ_ has on *β*_Γ_ is likely to inform the effect that varying either *µ*_Γ_ or *α*_Γ_ has on *β*_*D*_.

Finally, a positive correlation between *µ*_Γ_ and *T*_*D*_ is expected, as they both represent a form of average. For large *α*_Γ_, the gamma distribution becomes symmetric, hence *T*_*D*_ → *µ*_Γ_. However, the correlation becomes less linear as *α*_Γ_ decreases. In this case, *µ*_Γ_ − *T*_*D*_ and equivalently the skewness (defined by 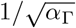 for the gamma distribution) increases (Fig. 4c).

## Discussion

This paper brings attention to and provides solutions to two distinct issues involved in modelling incubation periods of diseases. First, we derive a new model for delays between key events in an individual’s infection history, specifically the incubation period, that has justifiable mechanistic reasons for its validity. Second, we adapt methods for using incubation-period data, that is given as an integer number of days and has issues with bias, to fit models.

We considered the probability of an individual changing from the not-yet-symptomatic population to symptomatic for deriving our mathematical model. This approach led to obtaining a differential equation equivalent to the equation defining the exponential c.d.f. with a time-varying rate parameter. We then extended the model with further assumptions to further develop the differential equation describing the incubation period. We considered the likely event that the probability of symptom onset after infection is proportional to the bacterial load before saturating at some large load, as well as considering that bacterial population is expected to grow exponentially. Further, we derived a specific distribution within the Burr family that satisfies a Markovian property of long incubation periods. Other trial functions for *G*(*t*) may offer results at least as good as this new model, and some in-host dynamics which affect the rate of symptom onset in populations could be considered for specific diseases to provide even more optimal forms of the Burr model.

Further, by considering models that account for the fact that both the infection and symptom onset times are not exactly known (doubly interval-censored models), we have obtained expected incubation periods that are statistically significantly less than previously thought (by a whole day) using standard statistical distributions with incubation-period data. The mathematical derivation of the new model and implementation of this model with doubly interval-censored methods address both these problems, as we arrive at a mechanistic model for incubation periods. Our model has few restrictions on which diseases it can be applied to, as well as highlights the need to account for the censored nature of the data due to statistically significant difference recorded in calculated incubation periods when incorporating the DI methods into the model.

Our argument leading to the Burr family of distributions provides a valid incubation-period model, but does not consider factors such as an individual’s age, levels of immune response, susceptibility, doses received or the disease-specific in-host dynamics at play which determine if and when an individual becomes ill with an infection. For example, frailty may mean faster onset of symptoms, as may higher doses. This flexibility means that the exact disease-specific in-host dynamics are not considered, and to derive a model considering the biological processes at play with a given disease, a different model would have to be derived based on the details of those dynamics.

In Figure 1 of S1 Appendix in the Supplementary Material, we noticed that all Burr distributions valid over (0, ∞), apart from type X, exhibited a Markovian property for long incubation periods. Consequently, we compared the results of using this model to the other Burr distributions to judge the validity of the Markovian assumption. The type X provides successful results outperforming the gamma in nearly all of the analysis (we obtain mixed results when fitting to other diseases). However, when compared to all of the other Burr distributions, type X performed the worst when fitting to the original Legionnaires’ disease dataset, the original Legionnaires’ disease dataset with doubly interval-censored methods and the other diseases with doubly interval-censored methods. Further, type X visually fits the worst to the Legionnaires’ disease data (Fig. 2). These consistent results support our Markovian assumption for long incubation periods, and indicate that, although non-Markovian Burr distributions provide better-performing models to the widely used gamma model, the Markovian Burr models provide a further improvement in terms of distributional modelling.

Our proposed model can be applied in a number of ways in epidemiology and infectious disease modelling. For example, a common area of research for person-to-person transmissible diseases, such as COVID-19, is to develop compartmental and time-since-infection models where the infectivity of inflicted individuals infecting susceptible individuals in a population is modelled. Typically an exponential distribution from the point in time at which they are infected is used for modelling. However this approach can be improved upon by considering that an individual will have an incubation period before they are infectious to others. This improvement can be achieved by taking the convolution of the Burr incubation-period model and the exponential infectious period to gain a more reliable model for infectivity, improving the overall reliability of these models. In this work, we have limited to time delay distributions with range of times that are strictly positive, as must be the case with the incubation period. Some epidemiological distributions, such as generation time, are not bound by this constraint and so care would be needed in application.

Furthermore, we may consider diseases that do not have a person-to-person transmissible property such as Legionnaires’ disease, which has been the focus of this research. Researchers typically track backwards from symptom onset date to predict source location of the infection for elimination and public safety. A more reliable model such as the model developed here can provide more accurate results when predicting locations or causes of Legionnaires’ disease cases, which will result in reduction of bacterial hot-spots and consequently cases of this disease.

This paper provides a flexible model that can reliably fit incubation-period data to a level that is not currently seen in the literature and is valid for a wide range of diseases. We have validated this with our results indicating that using the Burr family of distributions as a model for incubation periods are better performing than currently accepted models [4] for the diseases that we have analysed.

## Data Availability

All data produced in the present work are contained in the manuscript and referenced papers.

https://pubmed.ncbi.nlm.nih.gov/28669361/

https://bmcinfectdis.biomedcentral.com/articles/10.1186/s12879-018-3391-3

https://pubmed.ncbi.nlm.nih.gov/7973702/

https://pubmed.ncbi.nlm.nih.gov/21242803/

## Acknowledgements

NJ acknowledges support from the Engineering and Physical Sciences Research Council (EPSRC) and Mathematics and Data in Scientific and Industrial Modelling (MADSIM) at the University of Manchester for funding of their studentship.

IH was supported by the JUNIPER modelling consortium (grant MR/V038613/1) the National Core Study on Transmission (PROTECT) and by the UKRI Impact Acceleration Account (IAA 386). NJ and IH also acknowledge the UK Health Security Agency (UKHSA) for honorary contracts and funding (for IH). The views expressed are those of the author(s) and not necessarily those of the Department of Health or UKHSA.

## Supporting information

**S1 Appendix**. Moments calculations for derived Burr and scaled type XII distributions. Further Legionnaires’ disease modelling analysis of mean incubation period and cumulative hazards.

**S2 Figure**. Figures and parameter estimates of anthrax, campylobacteriosis and salmonellosis datasets with model fits for gamma, burr types III, X, XII and the derived Burr based on the original and doubly interval-censored methods.

**S3 Code**. R code for conducting analysis and producing plots in this research. https://github.com/NyallJamieson/Burr-Incubation-Period

**S4 Data**. Incubation period data for the diseases analysed in this research.

